# Resting-state network plasticity induced by music therapy after traumatic brain injury

**DOI:** 10.1101/2020.05.29.20116509

**Authors:** Noelia Martínez-Molina, Sini-Tuuli Siponkoski, Linda Kuusela, Sari Laitinen, Milla Holma, Mirja Ahlfors, Päivi Jordan-Kilkki, Katja Ala-Kauhaluoma, Susanna Melkas, Johanna Pekkola, Antoni Rodriguez-Fornells, Matti Laine, Aarne Ylinen, Pekka Rantanen, Sanna Koskinen, Benjamin Ultan Cowley, Teppo Särkämö

## Abstract

Traumatic brain injury (TBI) is characterized by a complex pattern of abnormalities in resting-state functional connectivity (rsFC) and network dysfunction, which can potentially be ameliorated by rehabilitation. In our previous randomized controlled trial, we found that a 3-month neurological music therapy intervention enhanced executive function (EF) and increased grey matter volume in the right inferior frontal gyrus (IFG) in patients with moderate-to-severe TBI (N=40). Extending this study, we performed longitudinal rsFC analyses of resting-state fMRI data using a ROI-to-ROI approach assessing within-network and between-network rsFC in the frontoparietal (FPN), dorsal attention (DAN), default mode (DMN), and salience (SAL) networks, which all have been associated with cognitive impairment after TBI. We also performed a seed-based connectivity analysis between the right IFG and whole-brain rsFC. The results showed that neurological music therapy increased the coupling between the FPN and DAN as well as between these networks and primary sensory networks. By contrast, the DMN was less connected with sensory networks after the intervention. Similarly, there was a shift towards a less connected state within the FPN and SAL networks, which are typically hyperconnected following TBI. Improvements in EF were correlated with rsFC within the FPN and between the DMN and sensorimotor networks. Finally, in the seed-based connectivity analysis, the right IFG showed increased rsFC with the right inferior parietal and left frontoparietal (Rolandic operculum) regions. Together, these results indicate that the rehabilitative effects of neurological music therapy after TBI are underpinned by a pattern of within- and between-network connectivity changes in cognitive networks as well as increased connectivity between frontal and parietal regions associated with music processing.

## 1. Introduction

Each year there are over 50 million cases of traumatic brain injury (TBI), and it has been estimated that approximately half of the world’s population will sustain at least minor TBIs during their lifetime [1]. The consequences of TBI can be fatal; it is the leading cause of mortality in young adults and a major cause of death and disability across all ages worldwide. Depending on the injury mechanism, TBI can cause different pathophysiological changes in the brain such as diffuse axonal injury (DAI), bleeding and contusions. Due to TBI’s widespread effects that are dominant in the white matter tracts, it can be viewed foremost as a disorder of large-scale intrinsic connectivity networks [2]. DAI has also been shown to correlate with persistent cognitive impairments after TBI [3]. Despite the broad variety of symptoms that can follow after TBI, the most prominent cognitive impairments affect attention, memory and executive function (EF) [1, 3-6]. These high-level cognitive functions require the integration of information across spatially distinct brain regions, which make them particularly vulnerable to connectivity problems. In fact, deficits in EF are deemed to be the core symptoms of TBI [7, 8], particularly in moderate-to-severe cases that are at focus here. Although there is no consensus on the exact definition of EF, it is thought to encompass several cognitive processes including the core of set shifting, inhibition, and updating [9]. Given the heterogeneous and complex nature of TBI, and the major burden imposed upon individuals and society, there is an urgent need to develop novel and motivating rehabilitation strategies that target multiple deficits simultaneously, yet with a primary focus on EF.

Music is a very promising tool in TBI rehabilitation, because both music listening and participating in musical activities evoke widespread brain activation [10]. Musical creativity has recently been linked to the functioning of various resting-state networks including the default-mode, executive, salience, limbic and motor-planning networks [11]. It has also been shown that musical training enhances EF in healthy subjects and increases the engagement of the cognitive control network, which shares most of its nodes with the frontoparietal network [12-20]. Crucially, music has been shown to be an effective tool in enhancing cognitive and emotional recovery in neurological patients [21-24]. Since brain injury patients are usually able to enjoy and participate in musical activities [25], neurological music therapy can potentially contribute to restore the EF deficits observed in TBI patients [26]. Until recently, this question had been addressed by only three studies exploring the cognitive effects of music-based interventions after TBI [27-29]. Evidence from these studies indicated that music-based rehabilitation can indeed lead to cognitive recovery after brain injury, especially in the domain of mental flexibility, as well as activity and connectivity changes involving the orbitofrontal cortex, whose damage after TBI is associated with behavioral impairment [30]. However, these studies presented important limitations with regard to the sample size, lack of a proper randomized controlled design including a patient control group, and inclusion of patients with brain injury not caused by trauma.

We have conducted the first-ever randomized controlled trial (RCT) of neurological music therapy in a cohort of 40 moderate-to-severe TBI patients, where different domains of EF, attention, and memory were systematically analyzed. We used a single-blind cross-over design with 3 time-points (baseline/3-months/6-months) for neuropsychological assessment and s/fMRI acquisition. The neurological music therapy consisted of 20 individual therapy sessions held by a trained music therapist over a 3-month period (see 2. Materials and methods section for more details) and was targeted primarily to the rehabilitation of EF, attention, and working memory. In a previous publication, we reported that the music-based intervention induced cognitive improvement in general EF performance as well as in set-shifting [31].

In addition to demonstrating this improvement in neuropsychological performance, in [31] we conducted a voxel-based morphometry (VBM) analysis to investigate the volumetric changes induced by the music therapy. This analysis was motivated by previous work showing that environmental enrichment, such as that provided by musical activities, can increase the cognitive reserve and promote adaptive structural neuroplasticity changes [32-34], including stroke patients [22]. Our VBM results indicated that TBI patients showed an increase in grey matter volume (GMV) in different brain regions involved in music processing and cognitive function after the intervention. A therapy-induced increase in GMV was seen especially in the right inferior frontal gyrus and was correlated with enhanced set shifting ability.

Resting-state functional connectivity (rsFC) is characterized by task-free spontaneous fluctuations in brain activity that occur synchronously across spatially distant regions [35]. These fluctuations can be measured with the blood-oxygen-level-dependent response at low frequencies (usually under 0.15 Hz) and are spatially organized in resting-state networks (RSNs) [36] that mirror activity evoked during cognitive tasks [37, 38]. Examining rsFC after TBI is an active area of investigation motivated by the fact that DAI, a common pathology reported in all severities of TBI [39, 40], damages axonal wiring that partly underlies functional connectivity across RSNs [3]. The loss of integration of information in large-scale brain networks, which ultimately impairs high-level cognitive function, makes the study of rsFC after brain injury especially relevant [36, 41-43].

rsFC studies of TBI patients have revealed both increases and decreases in network connectivity, including the default mode (DMN) and salience (SAL) networks as well as multiple sensory and cognitive networks across the spectrum of injury severity [44-49]. For example, decreased rsFC has been observed within five network pairs (DMN-basal ganglia, attention-sensorimotor, frontal-DMN, attention-frontal and sensorimotor-sensorimotor; [49]) and within the motor-striatal network, in contrast to increased connectivity within the right frontoparietal network (FPN) [47]. In several cases, these abnormalities correlated with post-concussive symptoms [44, 48] and cognitive impairment [50].

Despite the complex abnormalities of interactions within and between RSNs following TBI, it is possible to identify some distinctive patterns. Within networks, reduced rsFC within the nodes of the DMN predicts attentional impairment [41]. Such association could be driven in turn by damage to the cingulum bundle connecting the nodes of the DMN. The temporal coordination between networks, which is important for efficient high-level cognitive function, has also shown consistent abnormalities after TBI. According to an influential model of cognitive control [51], the switching from automatic to controlled behavior is mediated by the interaction between the SAL and DMN networks, including deactivation of the DMN to attend unexpected external events. Indeed, TBI patients exhibit a failure to appropriately deactivate the DMN, which is associated with impaired response inhibition to a stop signal [41].

The paradoxical yet well-documented finding that functional connectivity may increase secondary to TBI [44-46, 52-56] has given rise to the “hyperconnectivity” hypothesis to explain the evolution of brain network reorganization after neurological disruption [57]. In this context, hyperconnectivity is defined as enhanced functional connectivity in the number or strength of connections. It is thought to affect preferentially RSNs with high-degree nodes, also known as network hubs, such as the frontoparietal (FPN), DMN and SAL networks. Although this hyperconnected state may be adaptive in the short-term in order to re-establish network communication through network hubs, Hillary and Grafman [57] have argued that it may have negative consequences due to the chronic enhancement of brain resource utilization and increased metabolic stress. In support of this view, abnormal functional connectivity has been associated with increased self-reported fatigue, which is a highly common and debilitating symptom after TBI [58]. Over a longer period of time this hyperconnectivity may even lead to late pathological complications including Alzheimer’s disease [59, 60], where amyloid beta deposition has been linked to the neurodegeneration of posterior DMN hubs with high metabolic rate [61-63].

In the present study, we extend our previous findings by analyzing rsFC from a subset of moderate-to-severe TBI patients (see 2. Materials and methods section for more details) in this music therapy RCT [31]. We used a seed-to-target approach to analyze the reconfiguration of RSNs induced by the neurological music therapy, selecting seeds from the nodes of four key networks: the DMN, SAL, FPN and dorsal attention (DAN) networks. The current work is grounded in two main hypotheses: (1) the music-based intervention leads to enhanced coordination activity between attention and executive function (DAN, FPN)-supporting networks, and sensory networks that are engaged during the music therapy; and (2) the music-based intervention elicits reduced connectivity of nodes within the SAL, DMN and FPN networks. In addition, we examined the relationship between the changes in rsFC within and between networks, and the therapy-induced improvement in EF shown previously with neuropsychological testing [31]. We anticipated that the FPN would be less connected in TBI patients with better EF performance; as the hyperconnectivity hypothesis predicts. Lastly, in a similar vein to the approach adopted by Han et al. [64], we conducted an exploratory seed-to-voxel analysis to elucidate the link between whole-brain rsFC and GMV enhancements induced by the neurological music therapy.

## 2. Materials and methods

We conducted a single-blinded cross-over RCT (trial number: NCT01956136) with a 6-month follow-up phase. Upon recruitment, patients were randomly assigned to one of two groups: AB and BA. During the first 3-month period, the AB group received neurological music therapy (NMT) in addition to standard care, whereas the BA group received only standard care. During the second 3-month period, the BA group received the music therapy intervention and standard care and the AB group received only standard care. Baseline measurements were administered at time-point 1 (TP1) and follow-up measurements were conducted at the 3-month cross-over point (TP2) and at the 6-month completion point (TP3).

### 2.1. Subjects and study design

The subjects analyzed herein are a subset of our previous study [31]. In the RCT, 40 TBI patients were recruited through the Brain Injury Clinic of the Helsinki University Central Hospital (HUCH), Validia Rehabilitation Helsinki, and the Department of Neurology of Lohja Hospital during 2014–2017. The inclusion criteria were: (1) diagnosed [Statistical Classification of Diseases and Related Health Problems, 10th revision (ICD-10)] TBI fulfilling the criteria of at least moderate severity [Glasgow Coma Scale (GCS) score:*<*=12 and/or post-traumatic amnesia (PTA) *>*=24h]; (2) time since injury *<*=24 months at the time of recruitment; (3) cognitive symptoms caused by TBI (attention, executive function, memory); (4) no previous neurological or severe psychiatric illnesses or substance abuse; (5) age 16–60 years; (6) native Finnish speaking or bilingual with sufficient communication skills in Finnish; (7) living in the Helsinki-Uusimaa area; and (8) understanding the purpose of the study and being able to give an informed consent.

To ensure steady allocation to both groups across the trial, randomization was performed using an online random number generator (https://www.random.org/) by a person not involved in patient recruitment or assessments. The randomization was stratified for lesion laterality (left/right/bilateral). Within each of these three strata, the randomization was done in batches of two consecutive patients. The trial was conducted according to the Declaration of Helsinki and was consistent with good clinical practice and the applicable regulatory requirements. The trial protocol was approved by the Coordinating Ethics Committee of the Hospital District of Helsinki and Uusimaa (reference number 338/13/03/00/2012) and all subjects signed an informed consent.

Of the 40 TBI patients enrolled to the trial, there were three dropouts before TP2 and another three dropouts before TP3. The dropouts were mainly due to lack of energy and motivation. Of the remaining 34 patients, one was excluded from the analyses due to intensive self-implemented piano training, which was not part of the trial protocol, and 10 were excluded from the analyses due to lack of fMRI data owing to MRI contraindications of the patients preventing scanning, technical difficulties, and artifacts in fMRI. This yielded a total sample of 23 patients (AB: n =15, BA: n =8) for the present study. This is a subset of the 25 patients sample with structural MRI scans from our previous study [31]. 12 patients sustained a lesion within 6 months or less at the time of recruitment and 11 patients were in the chronic phase (more than 6 months since injury).

### 2.2. Intervention

The NMT intervention consisted of 20 individual therapy sessions (2 times/week, 60 min/session) held by a trained music therapist (authors S.L. and M.H.) at Validia Rehabilitation Helsinki. The intervention required no previous musical experience and was adaptable to different degrees of injury across TBI patients. The intervention focused on active musical production with different instruments (drums, piano). Each session included three 20 min modules: (1) rhythmical training (playing sequences of musical rhythms and coordinated bimanual movements on a djembe drum and on own body), (2) structured cognitive-motor training (playing musical exercises on a drum set with varying levels of movement elements and composition of drum pads), and (3) assisted music playing (learning to play own favorite songs on the piano with the help of the therapist and using a special musical notation system called Figure Note). The difficulty level of the exercises was initially adjusted, and then increased in a stepwise manner within and across the sessions, to meet the skill level and progression of the patient. Musical improvisation was also included in all modules and encouraged throughout the therapy to facilitate creative expression (for more details on the neurological music therapy see [31]).

### 2.3. Neuropsychological assessment

An extensive battery of neuropsychological tests was used to measure the potential cognitive outcomes of the music therapy intervention. The battery is fully described in our previous article [31]. Briefly, it comprised standard neuropsychological tests of general EF [Frontal Assessment Battery (FAB) [65]], reasoning ability [Wechsler Adult Intelligence Scale IV (WAIS-IV) / Similarities and Block design subtests [66]], and verbal memory [Wechsler Memory Scale III (WMS-III) / Word Lists I-II subtests [67] and WAIS-IV / Digit Span subtest [66]] as well as computerized tests measuring different EF subcomponents, including set shifting [Number-Letter Task (NLT) [68]], working memory updating [Auditory N-back Task [69]], inhibition [Simon Task [70]], and sustained attention [Sustained Attention to Response Task (SART) [71]]. In addition, a self-report questionnaire measuring the severity of executive deficits [Behavioral Rating Inventory of Executive Function – Adult version (BRIEF-A) [72]] was included.

For correlation analyses with the fMRI data, we utilized only the behavioral data from the general EF (FAB total score), set shifting (NLT switching cost errors), and self-reported executive deficits (BRIEF-A / Self-Monitor and Inhibition scales). These measures were selected as they showed a significant positive effect of the music therapy intervention in our previous publication, as well as in another publication currently under review.

### 2.4. MRI Data Acquisition

Patients underwent structural MRI (sMRI) and resting-state functional (rs-fMRI) scanning, using an 8-channel SENSE head coil, in a 3T Philips Achieva MRI scanner (Philips Medical Systems) of the HUS Helsinki Medical Imaging Center at Helsinki University Central Hospital. In sMRI, we acquired 2 structural images, using: (1) a magnetization-prepared rapid acquisition gradient echo sequence (MPRAGE, hereafter referred to as T1) [repetition time (TR) = 9.9 ms; echo time (TE) = 4.60 ms, flip angle (FA) = 8 °, field of view (FOV) = 24.0 × 24.0 cm; matrix = 272 × 272; 187 slices; slice thickness 0.88 mm with no gap] and (2) a fluid-attenuated inversion recovery (FLAIR) sequence [repetition time (TR) = 8 ms; echo time (TE) shortest; angle= 50 °; field of view (FOV) = 9.8 × 9.7 cm; matrix = 228 × 226; 300 slices; slice thickness 0.60 mm with no gap].

In rs-fMRI, images were acquired with a T2*-weighted image sequence (TR/TE = 2000/35 ms; FA=90°; FOV = 23.0 × 23.0 cm; matrix = 80 × 80, 31 slices; slice thickness 4 mm with 0.5mm gap, 145 brain volumes; total scan time of 4 min 50 sec). During the rs-fMRI acquisition, the patients were instructed to remain still, eyes open to a fixation cross to ensure vigilance.

### 2.5. rs-fMRI Preprocessing

Data were preprocessed using a standard pipeline in Statistical Parametric Mapping software (SPM12, Wellcome Department of Cognitive Neurology, University College London) running under Matlab Release 2018b (The MathWorks, Inc., Natick, MA). The preprocessing methods applied included slice timing correction, realignment, segmentation, normalization to the MNI template (final voxel size 1mm isotropic) and smoothing with a Gaussian kernel of 8 mm full width at half maximum (FWHM). Focal brain lesions were detected in 11 patients. As the presence of lesions may influence the normalization algorithm, cost function masks (CFM) were defined for these 11 lesioned patients, to achieve optimal normalization with no post-registration lesion shrinkage or out-of-brain distortion [73]. Binary masks of the lesioned areas were obtained by manually drawing, on a slice-by-slice basis, the precise boundaries of the lesion directly into the T1 image from the TP1 session with MRIcron May 2, 2016 release (https://www.nitrc.org/projects/mricron). Accuracy of the CFM was validated by an expert neuroradiologist (author J.P.) who assessed multiple modalities of neuroimaging data acquired at TP1 (MPRAGE, FLAIR). For a lesion overlap map of the patients with visible lesions see Supplementary Figure S1.

Next, within-subject T1 images from all time-points (TP1, TP2, TP3) were coregistered using the T1 images from TP1 as reference to ensure that they remained in spatial alignment with the T1 images and CFM from this acquisition. All images and CFM were oriented to the anterior commissure before this coregistration step. Unified segmentation with medium regularization was applied to the T1 images (masked with CFM for those patients with visible lesions on the T1); grey matter (GM), white matter (WM), and cerebrospinal fluid (CSF) probability maps were obtained for each individual [74]. This procedure is common practice in patients with lesions [75].

After preprocessing, outlier identification was performed using the Artifact Detection Tools (ART) toolbox (https://www.nitrc.org/projects/artifactdetect). We identified outliers as scans that exceeded 3 standard deviations in Z-scores from the global BOLD signal or with framewise displacement > 2 mm. Regressor files containing the list of outliers for each patient and session (as one vector per outlier time-point, each containing 1 for the corresponding outlier and 0s for all other time-points) were saved for the removal of outlier effects.

### 2.6. rs-fMRI Denoising

We used denoising in order to minimize the variability due to physiological, outlier, and residual subject-motion effects. The preprocessed output files from SPM and ART were imported into the CONN toolbox v(18.b) (https://www.nitrc.org/projects/conn), including the following subject- and session-specific files: T1 and rs-fMRI scans; segmented GM, WM and CSF images; 6 realignment parameters after rigid body motion correction; and the regressor files containing the outliers. We used the default denoising pipeline in the CONN toolbox which comprises two steps in consecutive order: (1) linear regression of potential confounding effects in the BOLD signal and (2) temporal band-pass filtering. Step (1) implements an anatomical component-based noise correction procedure (aCompCor), including the following: noise components from cerebral white matter and cerebrospinal areas [76]; estimated subject-motion parameters plus their first-order derivatives [77]; identified outlier scans [78]; constant and first-order linear session effects [79]. After evaluation of the denoised outputs (residual BOLD time series), the number of noise components extracted from the white matter was increased to 10. In step (2), frequencies below 0.01 Hz were removed from the BOLD signal in order to reduce the slow signal drift (0.01 Hz < *f* < Inf).

### 2.7. Connectivity pattern analysis within and between large-scale networks

To further understand functional connectivity (FC) changes induced by the NMT from the perspective of large-scale networks, we used the 8 resting-state networks (RSNs) comprising 32 regions of interests (ROIs) or nodes in the CONN toolbox, which are derived from an independent component analysis (ICA) of 498 subjects from the Human Connectome Project [80]. The 8 RSNs included: default mode (DMN), salience (SAL), frontoparietal (FPN), dorsal attention (DAN), sensorimotor (SM), language (LAN), visual (VIS), and cerebellar (CER) networks. All of these networks have been widely reported in the resting-state literature.

We identified changes in both between-network and within-network connectivity from those networks that have shown alterations after TBI [3] or are associated with EF, memory and attention, which are the most commonly affected cognitive domains in TBI patients [3-6]. In particular, we selected ROIs from the DMN, SAL, FPN and DAN networks to examine the FC within their nodes and between their nodes and the rest of the RSN included in the CONN toolbox.

We used the Fisher-transformed bivariate correlation coefficient between a pair of ROIs time series [80]. Each pair of ROIs (i,j) constitutes an element in the ROI-to-ROI connectivity (RRC) matrix that can be calculated as follows:

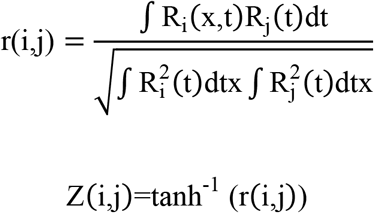

where R is the BOLD time series within each ROI, r is a matrix of correlation coefficients, and Z is the RRC symmetric matrix of Fisher-transformed correlation coefficients.

### 2.8. Longitudinal analysis of seed-based connectivity associated with changes in GMV

Several studies have shown that structural and brain connectivity changes may serve as biomarkers of plasticity in adults with chronic TBI [81, 82]. In our previous study, we demonstrated that neurological music therapy was able to induce morphometric changes in chronic TBI patients [31]. In particular, our findings indicated that GMV in the right inferior frontal gyrus (IFG), among other regions, increased significantly in AB and BA groups during the intervention versus control period as well as before and after the intervention period [31]. In addition, the increase in GMV in the right IFG correlated with cognitive improvement in the EF domain of set shifting [31].

In the current study, we selected the statistically significant clusters in the right IFG from the intervention period analysis to assess their seed-based connectivity in resting-state, because it was the only region to significantly correlate with therapy-induced EF improvement. The seeds included two clusters in the right IFG triangular part in standard space, that were imported into the CONN toolbox to compute seed-to-voxel functional connectivity. For more information regarding peak coordinate and size of the seeds see Table 4 in our previous study [31].

Seed-based connectivity (SBC) maps [80] between each seed and every voxel in the brain were obtained for each subject and session. SBC maps were computed as the Fisher-transformed Pearson’s bivariate correlation coefficients between a ROI BOLD time series and the BOLD time series of each individual voxel, using the equation:

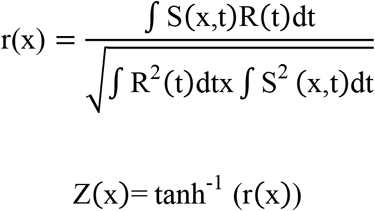

where S is the BOLD time series at each voxel, R is the average BOLD time series within a ROI, r is the spatial map of Pearson correlation coefficients, and Z is the SBC map of Fisher-transformed correlation coefficients for this ROI.

### 2.9. Quality assurance

We visually inspected all structural and functional MRI scans to ensure that patients had no significant brain atrophy. In rs-fMRI preprocessing, the quality of the preprocessed data was visually inspected using the quality assurance (QA) plots available in the CONN toolbox (QA normalization, registration, segmentation and framewise displacement). In rs-fMRI denoising, we inspected the distribution of connectivity values for each patient and session after denoising, and adjusted the number of noise components extracted from white matter to ensure that this distribution peaked around 0.

### 2.10. Group-level analysis

All group-level analyses were carried out in the CONN toolbox using the General Linear Model framework.

In the ROI-to-ROI (R2R) analysis, we calculated Group x Time interactions (AB>BA x TP2>TP1; BA>AB x TP3>TP2) using a mixed-model ANOVA. In order to increase the statistical power, we also performed a paired T-test pooling data across groups (pre- and post-intervention effect for the AB and BA groups; AB: T2>TP1 and BA: TP3>TP2). Furthermore, a paired T-test was performed during the control period (AB: T3>TP2 and BA: TP2>TP1) to ensure that the changes in rsFC were specifically caused by the intervention.

For the AB group, the denoised ROI time series from TP1 were entered as the pre-intervention condition and the denoised ROI time series from TP2 were entered as the post-intervention condition. For the BA group, the denoised ROI time series from TP2 were entered as the pre-intervention condition and the denoised ROI time series from TP3 were entered as the post-intervention condition. For the within-networks analysis, we examined the R2R connectivity of the ROIs within each of the selected four RSNs (DMN, SAL, FPN and DAN). For the between-network analysis, we examined the connectivity between the ROIs of the each of these RSNs (DMN, SAL, FPN, DAN) with every other RSN included in the CONN toolbox. As outlined above, we additionally performed a paired T-test during the control period for all the within- and between-network analyses. In this case, the denoised ROI time series from TP2 and TP3 were selected for the AB group and from TP1 and TP2 for the BA group.

In the SBC analysis, we performed a paired T-test comparing pre-versus post-intervention rsFC as the seeds were derived from two right IFG clusters obtained during the intervention period in our previous VBM analysis [31]. For the AB group, the beta maps from TP1 were entered as the pre-intervention condition and the beta maps from TP2 were entered as the post-intervention condition. For the BA group, the beta maps from TP2 were entered as the pre-intervention condition and the beta maps from TP3 were entered as the post-intervention condition. As in the R2R analysis, we additionally performed a paired T-test during the control period. In this case, the beta maps from TP2 and TP3 were selected for the AB group and from TP1 and TP2 for the BA group.

In addition, we confirmed that there were no differences in rsFC between the AB and BA groups at baseline that could be driven these effects. Specifically, we performed a two-sample T-test for the ROI-to-ROI analysis and the seed-to-voxel connectivity analysis. The ROIs and seeds were selected as described in 2.7 and 2.8 and we did the same connectivity comparisons as in the analyses described above.

The statistical parametric maps obtained were corrected for multiple comparisons based on Gaussian Random Field Theory [83]. The maps were first thresholded using a “height” threshold of p <0.001 and the resulting clusters thresholded with a cluster-level False Discovery Rate (FDR) -corrected p-value of p < 0.05 [84].

### 2.11. Resting-state functional connectivity and cognitive performance

We investigated the link between rsFC and cognitive performance improvements after therapy. We also performed Pearson’s bivariate correlation analyses between, on one hand, ROI-to-ROI (R2R) FC changes (found significant in the main analyses), and on the other hand, neuropsychological tests measuring general EF (FAB total score), set shifting (NLT switching cost errors), and the Self-Monitor and Inhibition subscales of BRIEF-A. The chosen performance measures were those which showed a statistically significant improvement after the music therapy [described in [31] and in an article under review]. We used the increment between pre- and post-intervention for the correlation with the intra-network FPN connectivity results and the increment between time-point 1 and 2 for the inter-network DMN connectivity results. FDR with the Benjamini & Hochberg method [85] was applied to the number of behavioral tests to correct for multiple comparisons.

## 3. Results

In this rs-fMRI longitudinal study, 145 brain volumes from 23 patients (AB group, n=15; BA group, n=8) were acquired at each of the three time-points: TP1, (baseline), TP2 (3-months) and TP3 (6-months).

### 3.1. Demographics

The demographic characteristics of the cohort are summarized in Table 1. As in our previous publication [31], we confirmed that there were no differences in sociodemographic information, clinical factors or musical background in the current sample of patients.

**Table 1.** Demographic, clinical and musical background information (N=23 TBI patients).

|  | All | AB | BA | Difference between groups (p) |
| --- | --- | --- | --- | --- |
| **Demographic information** |  |  |  |  |
| **Age** | 41.4 (13.2) | 41.1 (14.4) | 42.1 (11.6) | .860 (t) |
| **Gender** (female / male) | 10 / 13 | 7 / 8 | 3 / 5 | 1.000 (F) |
| **Handedness** (right/left/both) | 22 / 0 / 1 | 14 / 0 / 1 | 8 / 0 / 0 | 1.000 (F) |
| **Education in years** | 14.4 (2.5) | 14.0 (2.6) | 14.9 (2.2) | .420 (t) |
| **Clinical information** |  |  |  |  |
| **GCS** | 12.9 (3.5) | 12.8 (3.6) | 13.0 (3.6) | .967 (U) |
| **PTA classification<sup>a</sup>** | 1.8 (1.0) | 1.7 (1.1) | 1.9 (0.9) | .740 (t) |
| **Cause of injury** (traffic-related / fall / other) | 7 / 12 / 4 | 5 / 9 / 1 | 2 / 3 / 3 | .222 (F) |
| **Time since injury** (months) | 7.8 (6.1) | 8.0 (6.1) | 7.4 (6.5) | .820 (t) |
| **Lesion laterality<sup>b</sup>** (left / right / both) | 6 / 1 / 12 | 3 / 1 / 9 | 3 / 0 / 3 | .495 (F) |
| **DAI<sup>c</sup>** (yes / no) | 11 / 11 | 6 / 8 | 5 / 3 | .659 (F) |
| **Hemorrhages, bleeds or ischemic injury<sup>c</sup>** (yes / no) | 13 / 9 | 9 / 5 | 4 / 4 | .662 (F) |
| **GOSE<sup>c</sup>** | 5.4 (1.4) | 5.2 (1.5) | 5.6 (1.2) | .521 (t) |
| **NOS-TBI<sup>d</sup>** | 2.0 (2.2) | 2.1 (2.2) | 1.9 (2.5) | .848 (t) |
| **Musical background** |  |  |  |  |
| **Instrument playing** (yes / no) | 15 / 8 | 10 / 5 | 5 / 3 | 1.000 (F) |
| **Years of playing** | 4.7 (10.4) | 5.0 (11.7) | 4.0 (6.5) | .823 (U) |
| **Singing** (yes / no) | 11 / 12 | 8 / 7 | 3 / 5 | .667 (F) |
| **Years of singing** | 5.8 (11.7) | 8.2 (14.1) | 1.5 (3.1) | .330 (U) |
| **Dancing** (yes/no) | 11 / 12 | 8 / 7 | 3 / 5 | .667 (F) |
| **Years of dancing** | 5.3 (10.7) | 5.5 (11.3) | 4.9 (10.4) | .714 (U) |

<sup>a</sup> 1=mild (< 24 hours); 2=moderate (1-7 days); 3=severe (>7days); 4=very severe (>4 week); <sup>b</sup> Based on MRI-findings; <sup>c</sup> Glasgow Outcome Scale Extended; <sup>d</sup> Neurological Outcome Scale for TBI.

### 3.2. Between-network and within-network resting-state functional connectivity

To investigate the positive effect of the neurological music therapy on the coordinated activity between large-scale resting-state networks (RSNs) that are important for high-level cognitive function, we performed a ROI-to-ROI (R2R) analysis focusing primarily on the frontoparietal (FPN), dorsal attention (DAN), salience (SAL) and default mode (DMN) networks as source ROIs. Figure 1 shows the results of the between-network connectivity for these four RSNs, in a paired T-test of pre-versus post-intervention pooling together data from AB and BA groups (Figure 1A-C) and in the Group by Time (AB>BA x TP2>TP1) interaction (Figure 1D) with p < 0.05 FDR-corrected at the seed level.

**Figure 1.**
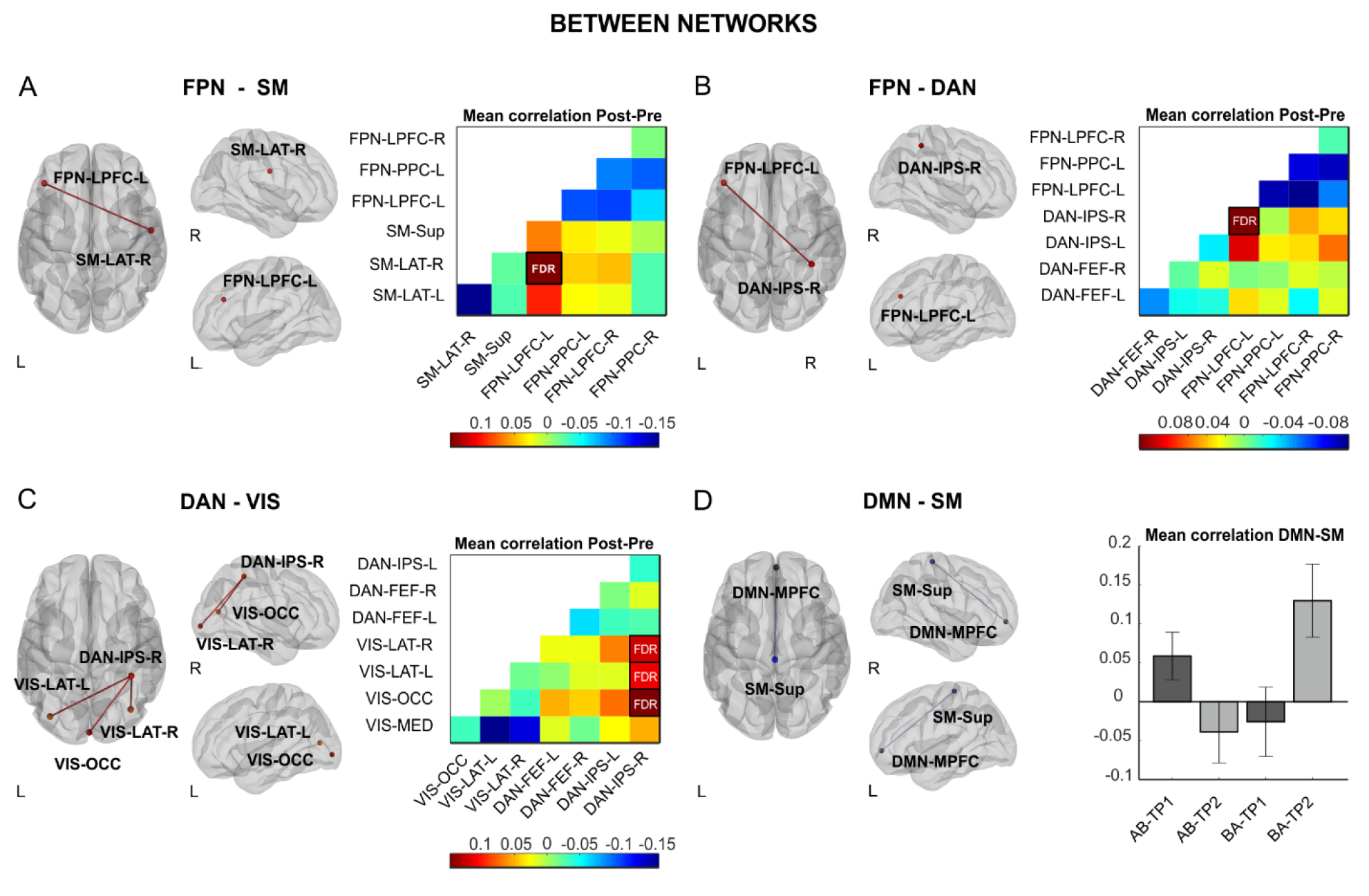
Changes in between-network connectivity induced by the neurological music therapy intervention. Nodes are overlaid on a rendered semitransparent brain generated using CONN. Connectivity matrices display the mean post-minus pre-intervention (A-C) Fisher-transformed Z-score correlation values for each node. The bar plots (D) show the effect size of the AB>BA and TP2>TP1 interaction represented by the Fisher-transformed Z-score correlation values for each node. *FPN: frontoparietal; SM: sensorimotor; DAN: dorsal attention; VIS: visual; DMN: default mode network; FEF: frontal eye field;, IPS: intraparietal sulcus; LPFC: lateral prefrontal cortex; LAT: lateral; MED: medial; MPFC: medial prefrontal cortex; OCC: occipital; PCC: posterior cingulate cortex; PPC: posterior parietal cortex; Sup: superior; R: right; L: left*.

In the pre-versus post-intervention comparison (see Table S1 for p and T values), we found that the FPN increased its temporal coupling with the sensorimotor (SM) and DAN after the music-based intervention. Likewise, when the DAN was used as a source, we observed increased between-network connectivity with several nodes of the visual (VIS) network induced by the intervention. In particular, the lateral prefrontal cortex in the left hemisphere from the FPN increased its functional connectivity with the right sensorimotor lateral node from the SM network (Figure 1A) and the right intraparietal sulcus from the DAN network (Figure 1B). In addition, the latter was also highly connected with the visual occipital and bilateral visual lateral nodes from the VIS network (Figure 1C). By contrast, in the AB>BA x TP2>TP1 interaction (Table S1), we found that the spontaneous fluctuations between the medial prefrontal cortex from the DMN network and the sensorimotor superior node from the SM network were less coordinated after the intervention (Figure 1D). We did not find any change in the between-network connectivity for the BA>AB x TP3>TP2 interaction.

When comparing AB and BA groups at baseline, we did not find any difference in the inter-network connectivity between the nodes of the FPN, DAN, SAL, DMN networks and every other node from the RSNs included in the CONN toolbox. To confirm that the pre-versus post-intervention changes in rsFC were driven by the music therapy, we also conducted a paired T-test pooling together data from the AB and BA groups for the time-points in which they were not participating in the intervention (control period). In this analysis, we found no significant between-network increases in rsFC for the FPN-SM, FPN-DAN and DAN-VIS pairs.

Next, we compared the connectivity within networks (Figure 2). In this case, only the pre-versus post-intervention comparison pooling together AB and BA groups (see Table S2 for p and T values) yielded significant results with p < 0.05 FDR-corrected at the seed level. The FPN and SAL networks showed a reduction in the functional connectivity between several of their constituent nodes. Specifically, the lateral prefrontal cortex node in the left hemisphere exhibited less coupling with its contralateral counterpart and with the posterior parietal cortex node in the left hemisphere (Figure 2A). Similarly, within the SAL network, the supramarginal gyrus node in the left hemisphere showed a decreased coupling with its contralateral counterpart and with the anterior insula node in the right hemisphere (Figure 2B).

**Figure 2.**
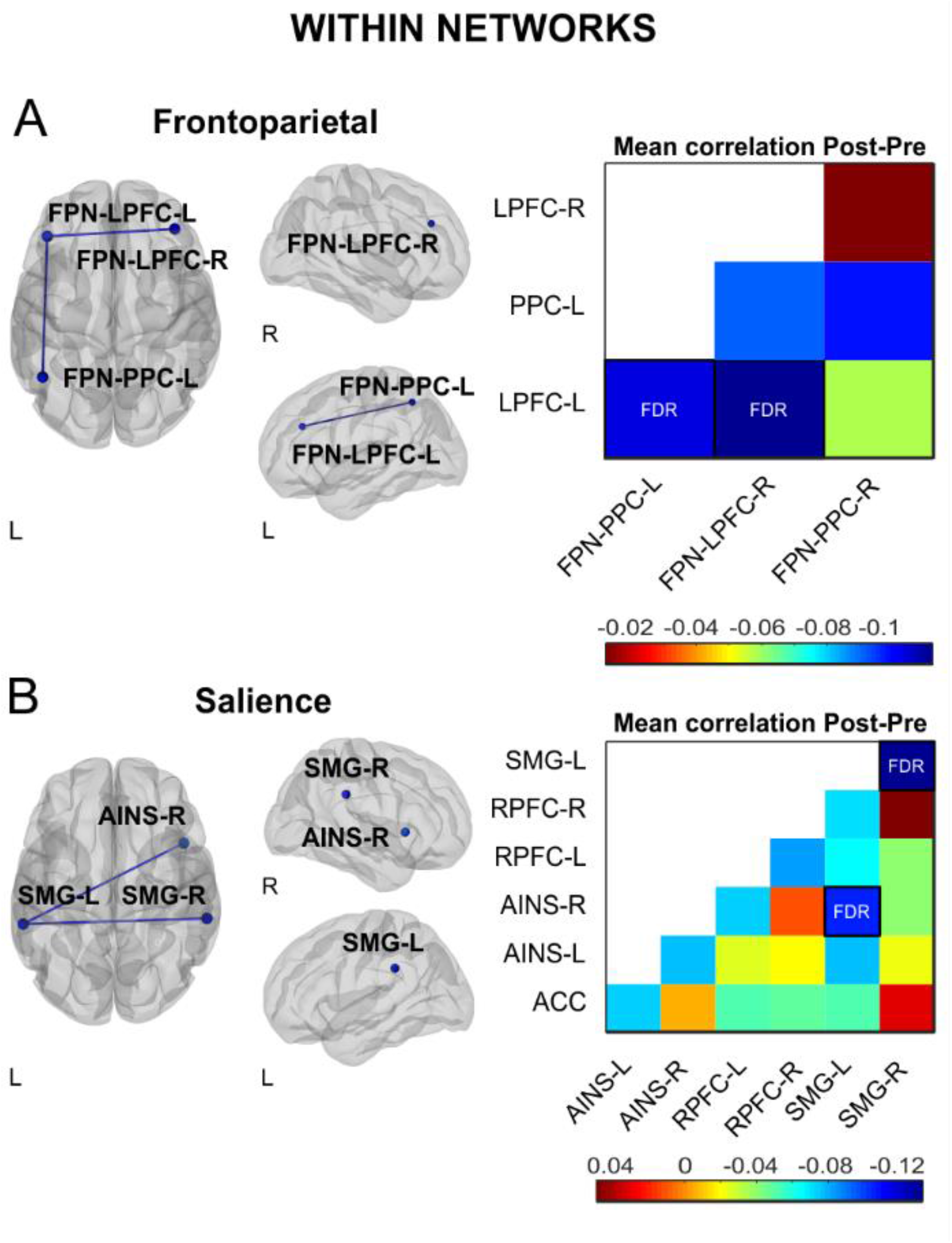
Changes in within-network connectivity induced by the neurological music therapy intervention. Nodes are overlaid on a rendered semitransparent brain generated using CONN. Connectivity matrices display the mean post-minus pre-intervention Fisher-transformed Z-score correlation values for each node. *AINS: anterior insula; IPS: intraparietal sulcus; LAT: lateral; LPFC: lateral prefrontal cortex; MPFC: medial prefrontal cortex; OCC: occipital; PPC: posterior parietal cortex; RPFC: rostral prefrontal cortex; SMG: supramarginal gyrus; Sup: superior; R: right; L: left*.

When comparing AB and BA groups at baseline, we did not find any difference in the intra-network connectivity of the RSNs included in the CONN toolbox. Furthermore, the within-network rsFC for the FPN, DAN, SAL and DMN networks did not change significantly during the control period.

### 3.3. Correlation between executive function and resting-state functional connectivity

Next, we investigated how the changes in RSNs induced by the neurological music therapy (see above) were associated with the parallel improvement in cognitive function, specifically in general EF (FAB score), set shifting (NLT switching cost errors), and self-reported executive deficits (BRIEF-A Self-Monitor and Inhibition subscales). We found statistically significant associations between the therapy-induced cognitive improvement and within-network and between-network changes in RSNs after the intervention (Figure 3). Decreased within-network FC in the left and right lateral prefrontal cortex nodes of the FPN correlated significantly with decreased BRIEF-A Self-Monitor scores (r = 0.485, p = 0.013, FDR-adjusted p = 0.039, Figure 3A, left) and showed a marginal trend with increased FAB scores (r = −0.372, p = 0.040, FDR-adjusted p = 0.060, Figure 3A, right). Decreased between-network FC between the DMN (medial prefrontal cortex node) and the SM (superior sensorimotor cortex node) networks correlated significantly with increased FAB scores (r = −0.559, p = 0.003, FDR-adjusted p = 0.009, Figure 3B, left) and showed a marginally significant with decreased NLT switching cost errors (r = 0.347, p = 0.052, FDR-adjusted p = 0.078, Figure 3B, right). There were no other significant correlations. Together, these results indicate that those patients who showed a larger reduction in connectivity within the FPN network and between the DMN and SM networks after training, exhibited greater improvement in general EF (higher FAB scores) and set shifting ability (less NLT errors), as well as greater reduction in executive deficits in self-monitoring (smaller BRIEF-A Self-Monitor scores). Thus, network-specific patterns of functional connectivity induced by the music-based intervention were associated with improvement in EF.

**Figure 3.**
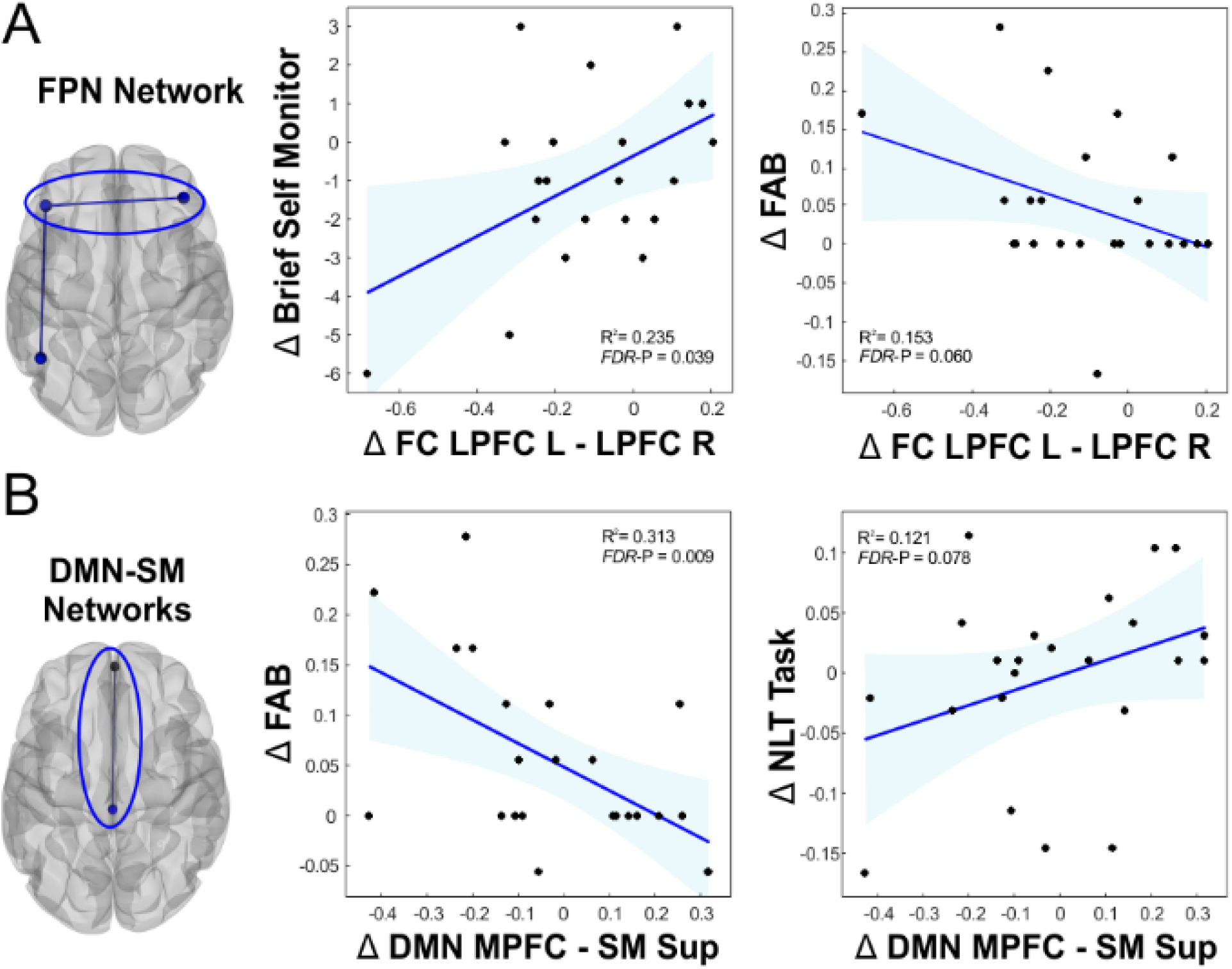
Within- and between-network functional connectivity changes associated with cognitive recovery induced by the neurological music therapy intervention. A: The behavior and functional connectivity values are derived from the pre-post intervention comparison, which was significant for the within-network connectivity changes in the FPN. B: The behavior and functional connectivity values are derived from the AB>BA and TP2>TP1 interaction, which was significant for the network connectivity changes between the DMN and SM network. The scatter plots represent the bivariate Pearson correlation and shaded areas represent the 95 % IC prediction bounds. *FPN: frontoparietal; DMN: default mode network; SM: sensorimotor; MPFC: medial prefrontal cortex; LPFC: lateral prefrontal cortex; Sup: superior; R: right; L: left*.

### 3.4. Seed-based resting-state functional connectivity

Previously, using voxel-based morphometry (VBM) we found that GMV in the right inferior frontal gyrus (IFG) increased significantly in both groups during the intervention period and that this was positively correlated with enhanced set shifting ability [31]. To examine the changes in resting-state functional connectivity (rsFC) associated with this increase in GMV, we performed seed-based connectivity (SBC) using as seeds the two clusters in the right IFG that resulted from our previous VBM analysis [31]. In the SBC analysis (results shown in Figure 4), a paired T-test of pre-versus post-intervention rsFC (pooling together AB and BA groups) revealed that during the intervention period the right IFG increased its connectivity with the right inferior parietal lobule (peak voxel x = 38, y = −46, z = 48; T= 4.95; Figure 4A) and the left Rolandic operculum (peak voxel x = −54, y = −2, z = 4; T= 6.07; Figure 4B) with p < 0.05 after FDR-correction at cluster level. There were no other significant clusters.

**Figure 4.**
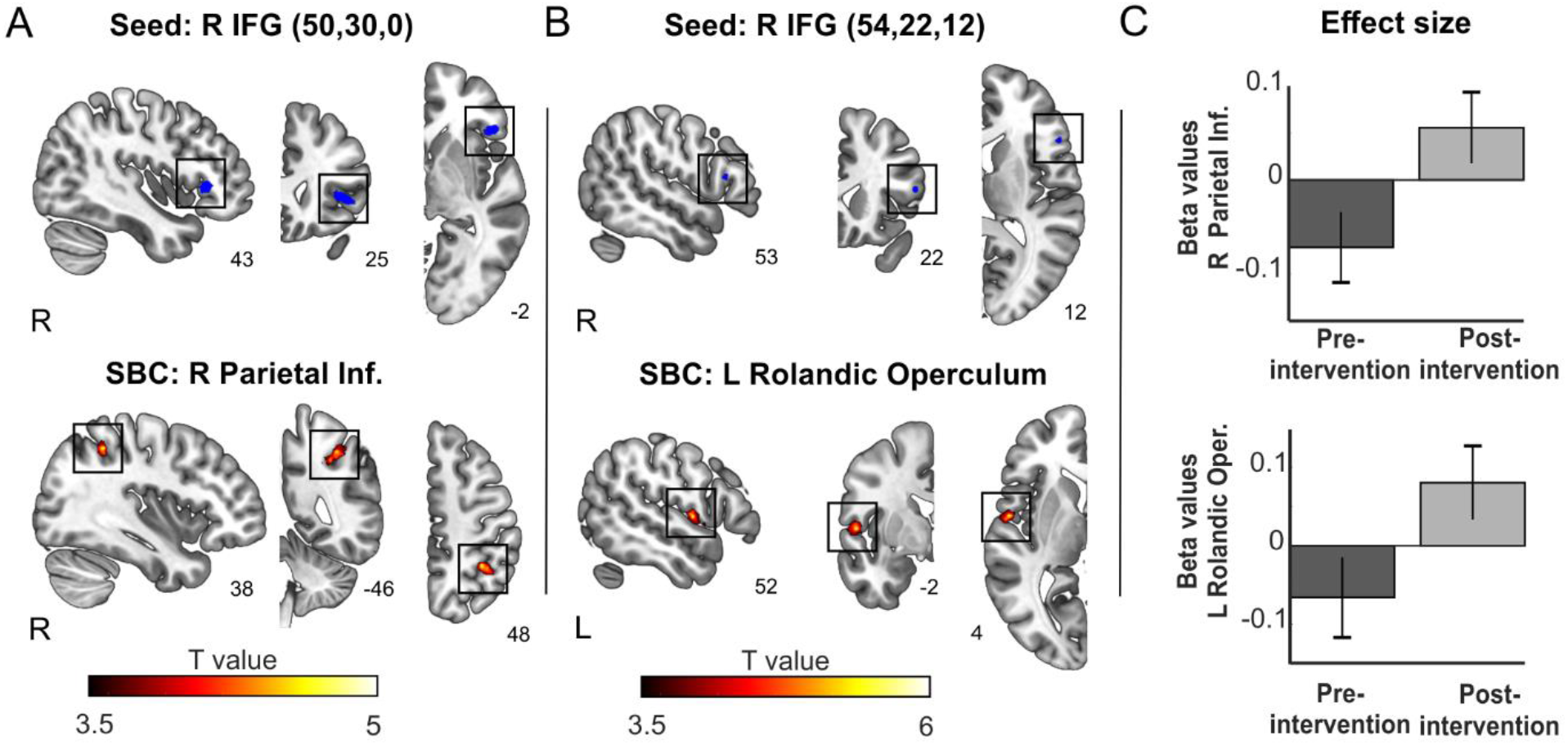
Changes in seed-based connectivity induced by the neurological music therapy intervention. A. *Top*: R IFG mask used as seed with peak coordinate x=50, y=30, y=0, overlaid on the Montreal Neurological Institute (MNI) template. *Bottom*: Statistical map showing the p < 0.05 FDR-corrected cluster in the R parietal Inf. B. *Top*: R IFG mask used as seed with peak coordinate x=54, y=22, y=12, overlaid on the MNI template. *Bottom*: Statistical map showing the p < 0.05 FDR-corrected cluster in the L Rolandic Operculum. C. Effect size plots displaying the functional connectivity (mean beta values) of each significant cluster in the pre-intervention relative to the post-intervention period. *IFG: Inferior Frontal Gyrus; Parietal Inf*.: *inferior parietal lobule, R: right; L: left*.

When comparing AB and BA groups at baseline, we did not find any difference in the whole-brain connectivity of the right IFG seeds. Moreover, there were no significant clusters in the SBC analysis with these right IFG seeds during the control period.

## 4. Discussion

We used resting-state functional connectivity (rsFC) methods to examine brain network changes induced by neurological music therapy (NMT) after moderate-to-severe TBI. We conducted a ROI-to-ROI approach with nodes from four networks [default mode (DMN), salience (SAL), frontoparietal (FPN) and dorsal attention (DAN)] as seeds and all network ROIs included in the CONN toolbox as targets. This analysis showed that music therapy strengthened network connectivity between FPN and DAN, and between these networks and the sensorimotor (SM) and visual (VIS) networks, respectively. In contrast, the music therapy intervention reduced the connectivity between the nodes of DMN and SM network. The within-network connectivity revealed specific nodes of the FPN and SAL wherein coupling decreased in the pre-versus post-intervention comparison. Importantly, the decrease in the FPN and DMN-SM connectivity was paralleled by cognitive improvement in executive function (EF). Finally, using a seed-based connectivity analysis, we demonstrated that the right inferior frontal gyrus (IFG), in which we previously observed increased grey matter volume (GMV) induced by the music therapy intervention [31], showed high connectivity with left frontal and right parietal regions, implicated in music processing [for a review, see [86]]. These results reflect substantial functional neuroplasticity changes in resting-state networks, which taken together might be interpreted as a shift from a hyperconnected state as reported in the TBI literature [57] to a connectivity state where efficient communication is maintained without a compensatory mechanism. We suggest that this represents a shift in executive functioning from a compensatory increase in cognitive control and attentional mechanisms to an optimal sensory-integrative functioning resembling that prior to the network disruption induced by the injury.

### 4.1. Changes in cross-modal integration

Our hypothesis (1) was that NMT enhances the rsFC between primary sensory networks and higher-order networks as a consequence of the iterated interaction between the sensory-cognitive systems recruited by music production, and perception [87]. This effect would be more prominent in the sensorimotor network as the percussion and piano training involved extensive fine and gross motor bimanual coordination, respectively. In addition, the assisted piano playing included music reading on a simplified notation system and therefore implied a high level of integration of information from the visual network.

The hypothesis (1) was conceptually inspired by the “global workspace” (or cross-modal integration) notion empirically supported by the convergence of unimodal sensory networks into cortical hubs that contribute to perceptual integration in the human brain [88]. Our results confirmed this hypothesis as we detected an increased coupling between the FPN and SM networks. The stepwise functional connectivity performed by Sepulcre et al. [88] highlighted the dorsolateral prefrontal cortex, which anatomically corresponds to the lateral prefrontal cortex node in the FPN, as one of the cortical hubs reached by seeds in the primary somatosensory cortex. Similarly, the intraparietal sulcus (IPS) node in the DAN was more connected with occipital nodes in the VIS network. This is not surprising given that visual streams have previously been postulated to converge in the IPS [89]. We also found an increase in rsFC between the FPN and DAN, which may be related to improved regulation of perceptual attention as recent work using meta-analytic tools has revealed [90].

### 4.2. Reduced connectivity in network hubs

In hypothesis (2) we proposed that the DMN, SAL and FPN connectivity would be downregulated by the NMT. Our reasoning was that the repetitive engagement of cognitive control and EF during the intervention [18, 20] would counter the increased connectivity and cognitive load associated with TBI. Brain injury has a detrimental effect on automatic processes and increases the supervisory demand for the integration of information at all levels [91], leading to the symptoms of fatigue commonly reported by these patients [58]. The need to compensate for the excessive cognitive demand resulting from focal lesions and diffuse axonal injury is consistent with the hyperconnectivity hypothesis [54, 57, 92].

One prominent line of evidence in favor of the latter hypothesis comes from the observation that hyperconnectivity secondary to neurological damage, including TBI [52, 54, 56, 93], is centered around nodes with a hub connectivity profile. Network hubs can be defined in terms of their structural [94-97] or functional connectivity [88, 98-100], and are characterized by a high degree of connectivity with the rest of the brain, thus making a strong contribution to the global integration of information [101]. Although functional connectivity does not reflect direct anatomical connections, graph measures derived from both methodologies show great convergence with regards to the brain regions classified as hubs. These include regions overlapping with the nodes of the DMN, SAL and FPN such as the superior parietal and superior frontal cortex, the anterior and posterior cingulate cortex as well as anterior portions of the anterior insula [88]. According to the hyperconnectivity hypothesis, a major goal of the increase in functional connectivity following injury is to re-establish network communication through network hubs in order to maximize information transfer and minimize behavioral impairments [57].

Here, we found that the neurological music therapy decreased the functional connectivity within the FPN, involving a reduction in the bilateral communication between the lateral prefrontal cortex and between the lateral prefrontal cortex and the posterior parietal cortex in the left hemisphere. Considering the hyperconnectivity hypothesis, this finding accords with recent evidence indicating that the bilateral prefrontal cortices are the subcomponents of the FPN with the highest average degree at 3 and 6 months after moderate-to-severe TBI [54]. Several other studies reported a similar increase in connectivity in frontal regions and FPN, among mild TBI patients [45, 47, 52]. Our results also showed that the anterior insula in the right hemisphere was less connected with the supramarginal gyrus in the left hemisphere, and that interhemispheric connectivity in the latter was equally diminished after the intervention. This within-network connectivity reduction in the SAL network, including the anterior insula, is again consistent with the hyperconnectivity reported by Hillary et al. [54]. The increase in functional connectivity of the anterior insula salience network after TBI has also been supported by other longitudinal [41] and cross-sectional studies [44]. Regarding the DMN, our analysis revealed a decreased connectivity between the medial prefrontal cortex and the SM network. Given the interference that the activation of the DMN may exert on attentional switching [51], it may be the case that this reduced connectivity facilitates the sensorimotor coupling with other brain regions involved in music production and perception. The negative trend with EF would lend support to this idea (however, since this correlation was only marginally significant, it should be interpreted with caution).

One clinically-relevant implication from these findings is that the music therapy effectively targeted network hubs from the FPN and SAL networks, and hence may have contributed to the shift from a compensatory hyperconnected state to one that does not rely on enhanced connectivity for efficient network communication. Within the hyperconnectivity hypothesis framework, it has been argued that while this hyperconnectivity may be adaptive in the short-term, chronic hyperconnectivity may render network hubs vulnerable to late pathological complications due to the chronically increased metabolic stress. Although the exact mechanisms and cognitive consequences of hyperconnectivity remain to be elucidated, this proposal has found some support from longitudinal studies examining the temporal evolution of TBI recovery [54, 92]. For example, Roy et al. [92] performed a cost-efficiency analysis and found a peak in network strength in the frontal DMN and temporoparietal networks at 6 months post-injury, with some residual hyperconnectivity observable after one year but with diminished overall cost. In this context, the NMT could potentiate the cost-efficiency rebalancing in the TBI recovery trajectory. Though beyond the scope of this paper, this prediction could be directly tested by computing graph theory measures including cost, degree, as well as local and global efficiency.

Indirect support for this idea comes from our correlational results indicating improved performance in EF as a function of rsFC decrease within the FPN. The NMT was primarily designed to target a number of EFs (action planning and monitoring, inhibitory control, shifting), which are reflected by the three outcome measures (FAB, NLT, BRIEF-A) showing a correlation with the therapy-induced change in rsFC. In this context, it might be that the repetitive practice during the music intervention reduces the cognitive challenge and transfers to better executive functioning, thus reducing the need for a hyperconnected state to efficiently accomplish the task. This would fit well with the observation that the increased activity in the right prefrontal cortex and anterior insula cortex in a sample of TBI patients normalizes following practice of a working memory task [91].

### 4.3. Relationship between brain morphometry and rsFC changes

The simultaneous combination of both structural and rsFC measures in the context of rehabilitative strategies for TBI has only recently started to be explored [81]. However, it offers an interesting possibility since both brain morphometry [102] and rsFC [103] are malleable to musical training. By cross-linking results from our previous voxel-based morphometry publication with rsFC, we were able to identify changes in GMV co-occurring with whole-brain rsFC. We focused on the right IFG as this was the region showing a significant relationship between GMV increases and improved performance in set shifting [31]. Using two right IFG clusters from the pre-versus post-intervention comparison as seeds, we found enhanced connectivity between the right IFG and the left Rolandic operculum, which is known to be activated in the processing of music-evoked emotions [104]. The right IFG also showed therapy-induced enhanced rsFC with the right inferior parietal lobule (also known as Geshwind’s territory), which is particularly interesting since both regions are involved in musical syntax analysis [105-107] and have a direct anatomical connection via the anterior segment of the arcuate fasciculus [108]. Therefore, we can speculate that this effect could be mediated by a concomitant increase in the underlying structural connectivity. Future tractography studies in the same TBI cohort could specifically address this issue.

### 4.4. Limitations and future directions

This study has some limitations, which need to be considered when evaluating its findings. Although this is the largest RCT using neurological music therapy in moderate-to-severe TBI to date, the final sample size is relatively modest (N=23) and may preclude the detection of small effect sizes due to lack of statistical power. Given the difficulty in finding patients fulfilling all the inclusion criteria, we were more liberal in the selection of time since injury. For this reason, even if over half of our patients sustained TBI within 6 months or less at the time of recruitment, there is heterogeneity in our sample. While the interpretation of our findings is consistent with previous longitudinal studies examining rsFC in TBI, a direct comparison is not warranted and future RCT should consider a stratification of patients by time since injury. Another consideration is the cross-over design: this has the advantage that patients act as they own controls, but the potential risk of a carry-over effect for the group who first participated in the intervention. Regarding rsFC, we used the resting-state networks available in the CONN toolbox that were obtained using an ICA analyses (N=498) from the Human Connectome Project. This selection, however, limited the analysis of sensory-integrative network interactions since the auditory network was not included. As we have established that the music therapy effectively enhances these interactions, future analyses will benefit from including nodes from the auditory network based on brain parcellations [97] or ICA decomposition [36]. Physiological measures to account for the effect of breathing and heart rate in introducing structured noise were not acquired in the collection of this data set. Finally, although the human brain is a dynamic system, this study investigated how the neurological music affects the cross-modal integration across sensory and cognitive networks with a stationary FC approach by averaging time courses from seeds and targets. In order to gain a more fine-grained view of how the intervention influences network connectivity in the time scale, future work should examine differences in the dynamic FC between these segregated functional networks.

### 4.5. Conclusions

In conclusion, this study provides compelling evidence that NMT can lead to substantial functional neuroplasticity changes in resting-state networks after TBI. Our results lend support to the idea that the music intervention facilitated the integration of primary sensory information, by increasing the rsFC between multimodal and higher-level cognitive networks (FPN, DAN). The shift towards a less connected state within the FPN and SAL networks is also in line with the notion that chronic hyperconnectivity in network hubs after TBI can be maladaptive in the long-term. Importantly, we found a correlation between improved performance in EF and lesser connectivity in the FPN and the DMN-SM networks, which might be linked to reduced interference. Finally, the co-occurrence of changes in brain morphometry and rsFC connectivity, in the circuits supporting music processing, suggests a complex picture with interrelated plasticity across MRI modalities. Our findings suggest that rsFC changes in brain networks can serve as sensitive biomarkers for the efficacy of music-based rehabilitation after TBI. These findings are crucial from a clinical standpoint, as effective rehabilitative tools for reducing brain network dysfunction after TBI are still under development.

## Data Availability

This data has not been made available at a public repository yet.

## Data availability

The data that support the findings of this study are available from the corresponding author upon reasonable request.

## Supplementary information

The attached file of supplementary materials presents two tables with the results from the second-level analyses of the functional connectivity between and within networks and one figure showing the lesion overlap map. Table S1 shows the statistically significant results from the second-level analysis of functional connectivity between the nodes of the 4 resting-state networks of interest in this study (namely, Frontoparietal, Dorsal Attention, Salience and Default Mode networks) with every other resting-state network included in the CONN toolbox. Table S2 shows the statistically significant results from the second-level analysis of functional connectivity within the nodes of the 4 resting-state networks of interest in this study (namely, Frontoparietal, Dorsal Attention, Salience and Default Mode networks) included in the CONN toolbox. Figure S1 shows the lesion overlap map derived from 11 TBI patients with visible lesions.

## Preprint statement

A previous version of this manuscript has been made available in a preprint server: https://www.medrxiv.org/content/10.1101/2020.05.29.20116509v1?versioned=true.

## Acknowledgments

We would like to thank the multi-disciplinary team of experts participating in the planning and data collection: Music therapist Esa Ala-Ruona participated in planning of the intervention; Professor Mari Tervaniemi and neurologist Anne Vehmas took part in the initial planning of the study; Research nurse Veera Lotvonen recruited the participants with the help of neuropsychologists Jaana Sarajuuri and Titta Ilvonen. Above all we want to thank the patients with TBI and their family members who participated in the study.

## Supplementary Material

**Table S1.** Between-network connectivity results. This table shows the statistically significant results from the second-level analysis of functional connectivity between the nodes of the 4 resting-state networks of interest in this study (namely, Frontoparietal, Dorsal Attention, Salience and Default Mode networks) with every other resting-state network included in the CONN toolbox.

| **Pre- versus Post-Intervention Comparison (one-sided contrast)** | **T(22)** | **p-unc** | **p-FDR** |
| --- | --- | --- | --- |
| Frontoparietal-Lateral Prefrontal Cortex-Left AND Sensorimotor-Lateral-Right | 4.02 | 0.0003 | 0.0017 |
| Frontoparietal-Lateral Prefrontal Cortex-Left AND Dorsal Attention-Intraparietal Sulcus-Right | 3.21 | 0.0020 | 0.0140 |
| Dorsal Attention -Intraparietal Sulcus-Right AND Visual-Occipital | 2.97 | 0.0035 | 0.0245 |
| Dorsal Attention -Intraparietal Sulcus-Right AND Visual-Lateral-Right | 2.61 | 0.0080 | 0.0281 |
| Dorsal Attention -Intraparietal Sulcus-Right AND Visual-Lateral-Left | 2.39 | 0.0130 | 0.0303 |
| **AB>BA and TP2>TP1 Interaction (one-sided contrast)** | **T(22)** | **p-unc** | **p-FDR** |
| Default Mode Network-Medial Prefrontal Cortex AND Sensorimotor-Superior | -3.20 | 0.0021 | 0.0128 |

*p-unc = p-uncorrected; p-FDR= False Discovery Rate-adjusted p at the source level.*

**Table S2.** Within-network connectivity results. This table shows the statistically significant results from the second-level analysis of functional connectivity within the nodes of the 4 resting-state networks of interest in this study (namely, Frontoparietal, Dorsal Attention, Salience and Default Mode networks) included in the CONN toolbox.

| **Pre- versus Post-Intervention Comparison (one-sided contrast)** | **T(22)** | **p-unc** | **p-FDR** |
| --- | --- | --- | --- |
| Frontoparietal-Lateral Prefrontal Cortex-Left AND Frontoparietal-Lateral Prefrontal Cortex-Right | -2.58 | 0.0171 | 0.0494 |
| Frontoparietal-Lateral Prefrontal Cortex-Left AND Frontoparietal-Posterior Parietal Cortex-Left | -2.28 | 0.0329 | 0.0494 |
| Salience-Supramarginal Gyrus-Left AND Salience-Supramarginal Gyrus-Right | -3.02 | 0.0063 | 0.0322 |
| Salience-Supramarginal Gyrus-Left AND Salience-Anterior Insula-Right | -2.79 | 0.0107 | 0.0322 |

*p-unc = p-uncorrected; p-FDR= False Discovery Rate-adjusted p at the source level.*

**Figure S1.**
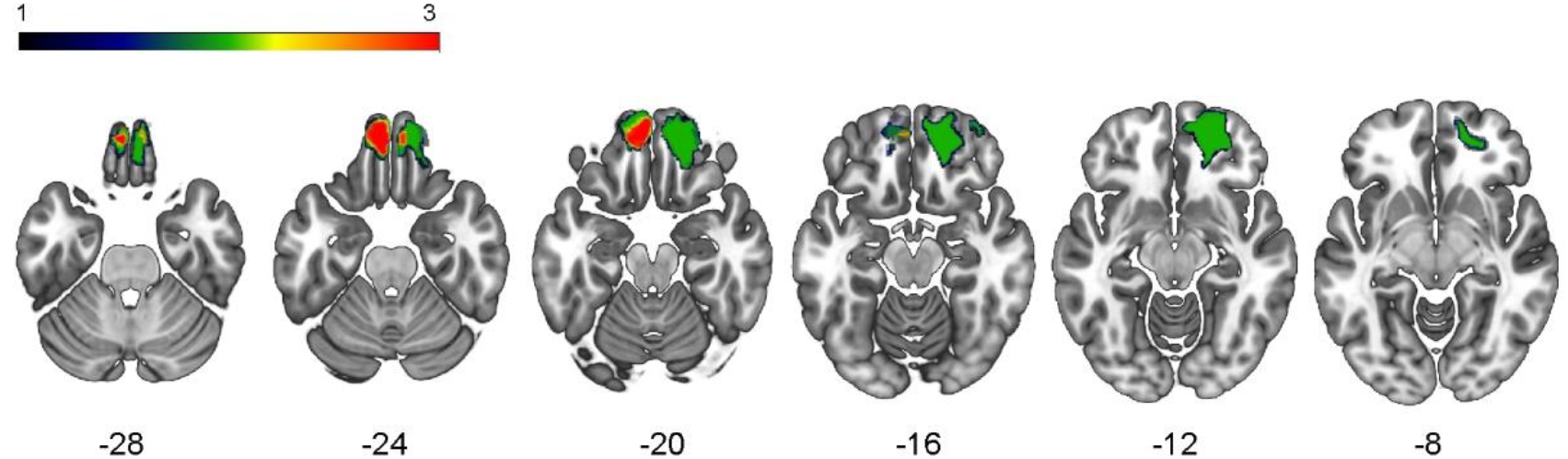
Lesion overlap of 11 traumatic injury patients with visible lesions. The colorbar indicates the number of patients with a lesion in each voxel (maximum 3 out of 11). Numbers below the axial slice denote the Z coordinate in mm.

